# Ultra-low DNA input into whole genome methylation assays and detection of oncogenic methylation & copy number variants in circulating tumour DNA

**DOI:** 10.1101/2020.02.11.20021964

**Authors:** Celina Whalley, Karl Payne, Enric Domingo, Andrew Blake, Susan Richman, Jill Brooks, Nikolaos Batis, Rachel Spruce, S-CORT Consortium, Hisham Mehanna, Paul Nankivell, Andrew D Beggs

## Abstract

**Background:** CpG methylation in cancer is ubiquitous and generally detected in tumour specimens using a variety of techniques at a resolution encompassing single CpG loci to genome wide. Analysis of samples with very low DNA inputs, such as formalin fixed (FFPE) biopsy specimens from clinical trials or circulating tumour DNA has been challenging and has only been typically at single CpG sites. Analysis of genome wide methylation in these specimens has been limited because of the relative expense of techniques need to carry this out. We present the results of low input experiments into the Illumina Infinium HD methylation assay on FFPE specimens and ctDNA samples.

**Methods:** For all experiments, the Infinium HD assay for Methylation was used. In total, forty-eight FFPE specimens were used at varying concentrations (lowest input 50ng), eighteen blood derived specimens (lowest input 10ng) and six matched ctDNA input (lowest input 10ng) / fresh tumour specimens (lowest input 250ng) were processed. Downstream analysis was performed in R/Bioconductor for QC metrics and differential methylation analysis as well as copy number calls.

**Results:** Correlation coefficients for CpG methylation at the probe level averaged R2=0.99 for blood derived samples and R2>0.96 for the FFPE samples. When matched ctDNA/fresh tumour samples were compared R2>0.91. Results of differential methylation analysis did not vary significantly by DNA input in either the blood or FFPE groups. There were differences seen in the ctDNA group as compared to their paired tumour sample, possibly because of enrichment for tumour material without contaminating normal. Copy number variants observed in the tumour were generally also seen in the paired ctDNA sample.

**Conclusions:** The Illumina Infinium HD methylation assay can robustly detect methylation across a range of sample types, including ctDNA, down to a input of 10ng. It can also reliably detect oncogenic methylation changes and copy number variants in ctDNA.

## BACKGROUND

Epigenetic change in the form of 5-methylcytostine is one of the most ubiquitous epigenetic changes in cancer genetics, controlling gene expression via repression of transcription factor binding due to its inaccessibility caused by methylation (1).

Methylation in cancer is of particular interest due to its association with specific phenotypes such as the CpG Island Methylator Phenotype (CIMP) that possess unique characteristics (2). In colorectal cancer, CIMP is associated with a hypermethylator phenotype (3), usually found in the right colon in female patients with advanced age, can be associated with microsatellite instability (due to DNA mismatch repair suppression) and has been suggested to be a biomarker for poor prognosis (4). In fact, the epigenome has been shown to vary widely across different cancer types and multiple methylation subtypes have been identified. The TCGA consortium identified four methylator subtypes associated with colorectal cancer, each associated with differing clinico-pathological characteristics (5). In head and neck squamous cell carcinoma (HNSCC), analysis of TCGA tumour methylation data has revealed both hyper- and hypomethylated gene sets for diagnostic purposes (6). In addition, 6 methylation clusters have been identified that correlate with clinicopathological characteristics such as HPV status, and also 3-year survival (7).

A particular focus of interest in study of the epigenome is in circulating tumour DNA (8) (ctDNA). This has attracted particular attention as a biomarker for disease, with the GRAIL consortium utilising a mutation based assay for circulating tumour DNA with over 90% accuracy for the detection of cancer (9). However, the use of methylation assays on this sample type has been restricted due to low input concentrations available from ctDNA. The UroMark consortium (10) has studied the use of bisulphite amplicon resequencing in the analysis of free urinary DNA. The DNA input required for this is relatively plentiful (> 30ng of urinary DNA) compared to that available for circulating tumour DNA (typically 10-20ng per 10mls of whole blood).

In order to develop new biomarker panels for the methylation based detection of disease, a whole genome approach is required. Several technologies exist for this purpose – whole genome bisulphite sequencing (11) (WGBS), reduced representation bisulphite sequencing (12) (RBBS) and methylation arrays, typically the Illumina Infinium assay (13). Both WGBS and RBBS require large input amounts of DNA (∼ 3ug) and are particularly expensive, the former especially so due to the increased depth of coverage required necessitating increased sequencing cost. Methylation arrays, the most popular of which is the Illumina Infinium assay require at a minimum, 200-300ng of bisulphite converted DNA, which can be from formalin fixed, paraffin embedded (FFPE) archival samples, repaired by use of a proprietary “DNA restore” kit. The input requirements, as specified by the manufacturer make this assay seemingly unusable for the requirements of low input and ctDNA experiements.

We hypothesised that as the Illumina Infinium assay requires a whole genome (14) amplification (WGA) step, the theoretical input to this kit could encompass concentrations seen in ctDNA samples. We therefore aimed to study the effect of input concentration of DNA on assay performance. As a secondary objective, we also attempted to develop an alternate, cheaper methodology for restoration of FFPE DNA in the Illumina Infinium assay in order to reduce costs.

## RESULTS

### Sample QC metrics

All samples across both blood, FFPE and fresh tissue passed QC metrics for their respective samples types. For samples C, 1 and 3 all had > 99% CpG detection across the range of DNA input (10-500ng). QC results are shown in tables 1 & 2. For FFPE samples from the SCORT consortium (n=16, across 50, 100 and 150ng input mass), all samples passed QC with > 95% CpG detection, reflecting the lower detection threshold provided by Illumina for lower quality samples.

**Table 1.** Shows CpG % detection levels from the fresh blood DNA samples processed on the Infinium® HumanMethylation 450k BeadChip. There are 485577 CpG sites on this array with the QC cut off for % CpG detection, at ≥ 99%.

| Sample ID | DNA input (ng) | Detected CpG<br>(0.05) | % CpG detection |
| --- | --- | --- | --- |
| C-10 | 10 | 483,570 | 99.59 |
| C-50 | 50 | 485,395 | 99.96 |
| C-100 | 100 | 485,475 | 99.98 |
| C-150 | 150 | 485,466 | 99.98 |
| C-200 | 200 | 485,473 | 99.98 |
| C-250 | 250 | 485,471 | 99.98 |
| C-300 | 300 | 485,455 | 99.97 |
| C-500 | 500 | 485,446 | 99.97 |
| 3-2_10 | 10 | 484,491 | 99.78 |
| 3-2_50 | 50 | 485,417 | 99.97 |
| 3-2_100 | 100 | 485,406 | 99.96 |
| 3-2_200 | 200 | 485,435 | 99.97 |
| 3-2_250 | 250 | 485,445 | 99.97 |
| 1-1_10 | 10 | 484,158 | 99.71 |
| 1-1_50 | 50 | 485,346 | 99.95 |
| 1-1_100 | 100 | 485,446 | 99.97 |
| 1-1_200 | 200 | 485,458 | 99.98 |
| 1-1_250 | 250 | 485,485 | 99.98 |
| 199-cfDNA | 10 | 457,792 | 94.28% |
| 207-cfDNA | 10 | 467,045 | 96.18% |
| 264-cfDNA | 10 | 480,752 | 99.01% |
| 268-cfDNA | 10 | 471,080 | 97.01% |
| 276-cfDNA | 10 | 471,041 | 97.01% |
| 288-cfDNA | 10 | 451,610 | 93.00% |
| 199-tissue | 250 | 485,046 | 99.89% |
| 207-tissue | 250 | 484,988 | 99.88% |
| 264-tissue | 250 | 484,727 | 99.82% |
| 268-tissue | 250 | 484,983 | 99.88% |
| 276-tissue | 250 | 484,442 | 99.77% |
| 288-tissue | 250 | 484,962 | 99.87% |

**Table 2:** Shows qPCR quality and CpG % detection levels per sample for each FFPE DNA input amount. There are 866,895 CpG sites on the Infinium® MethylationEPIC BeadChip arrays with a QC cut off of ≥90% detection with FFPE DNA.

| Sample ID | qPCR Delta<br>Cq value <5<br>(>5 poor) | % CpG<br>detection (p=<br>0.05) 150 ng<br>input | % CpG<br>detection (p=<br>0.05) 100 ng<br>input | % CpG<br>detection (p=<br>0.05) 50 ng<br>input |
| --- | --- | --- | --- | --- |
| SC00236A2 | 2.34 | 96.55 | 97.57 | 96.69 |
| SC00264A2 | 1.59 | 97.95 | 98.73 | 97.93 |
| SC00267A2 | 2.95 | 98.03 | 98.40 | 97.50 |
| SC00273A2 | 3.10 | 97.99 | 98.22 | 97.66 |
| SC00275A2 | 2.72 | 97.60 | 98.09 | 97.10 |
| SC00292A2 | 3.61 | 96.99 | 97.97 | 96.30 |
| SC00296A2 | 2.08 | 99.22 | 99.27 | 98.01 |
| SC00357A2 | 0.89 | 99.40 | 99.44 | 99.22 |
| SC00447A2 | 0.71 | 98.66 | 99.04 | 98.38 |
| SC00476A2 | 1.87 | 99.18 | 99.17 | 99.19 |
| SC00482A2 | 1.09 | 98.95 | 98.85 | 98.49 |
| SC00486A2 | 1.97 | 98.85 | 98.94 | 98.63 |
| SC00491A2 | 1.83 | 98.28 | 98.29 | 98.10 |
| SC00496A2 | 3.05 | 93.07 | 95.28 | 94.96 |
| SC00498A2 | 1.83 | 98.86 | 98.98 | 98.54 |
| SC00529A2 | 1.08 | 99.14 | 99.20 | 98.75 |

### Sample correlations

In order to understand whether there was a relationship between reducing input and loss of correlation between identical samples a correlation analysis was run using the *cor* command in the *psych* module of *R*.

For samples C, 1 and 3, all correlation coefficients were > 0.99 across all comparators (Pearsons p<0.001, Table 3). For the FFPE derived samples, the correlation coefficient was at least 96% in all samples. Correlation matrices and distributions for all sample types are shown in Figure 1. Correlation between sample inputs dispersed more widely as the input amount, with a particular widening around 10ng input across the blood derived samples. In the FFPE sample group these were more widely dispersed generally, however less of an effect in terms of dispersion at low input concentrations was seen. For the ctDNA samples, correlation was performed between all the samples, and lower correlations (typically < 0.95) and very wide dispersions were seen as the point of this plot was to demonstrate that they were not similar.

**Table 3:**
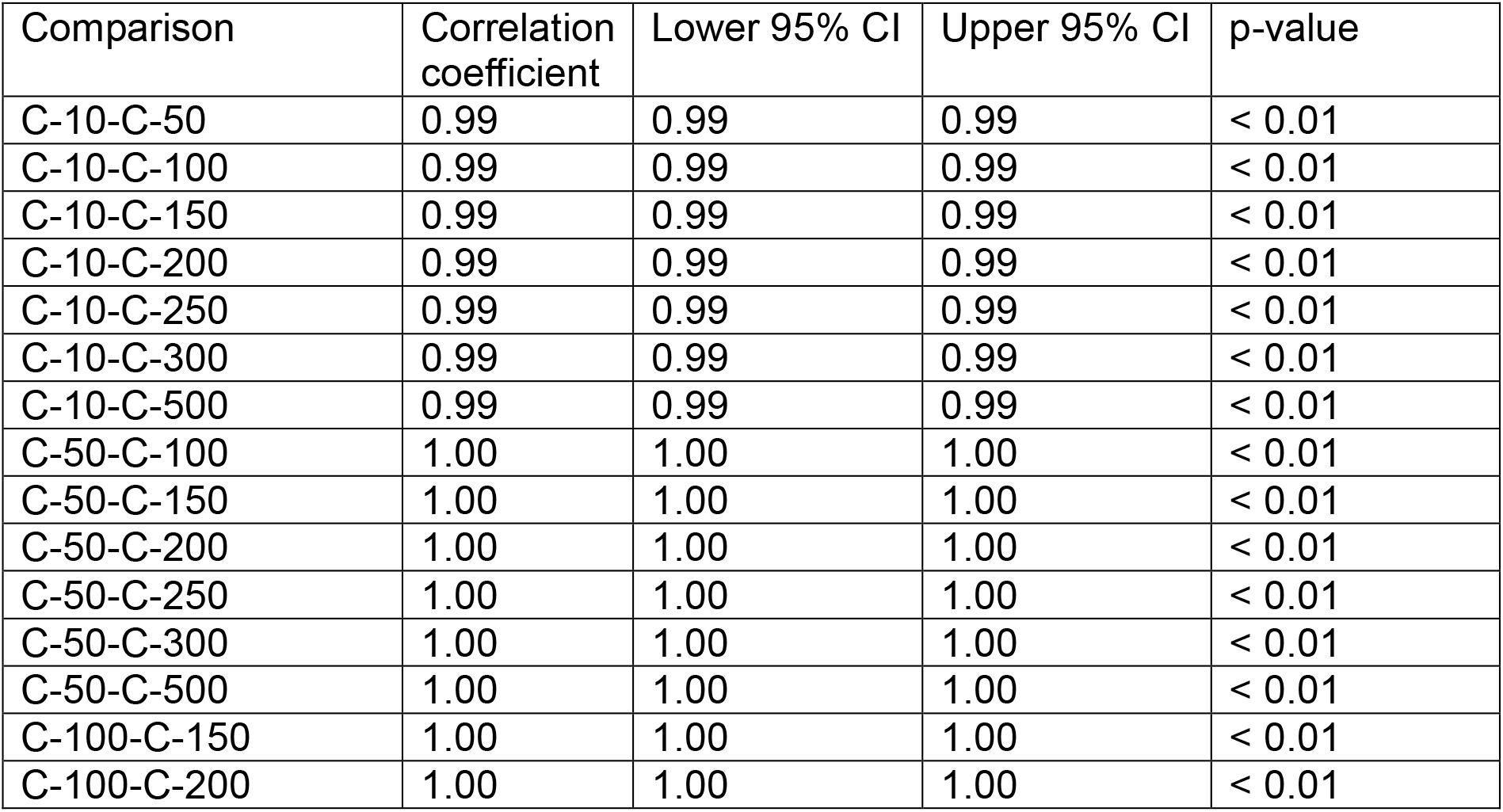
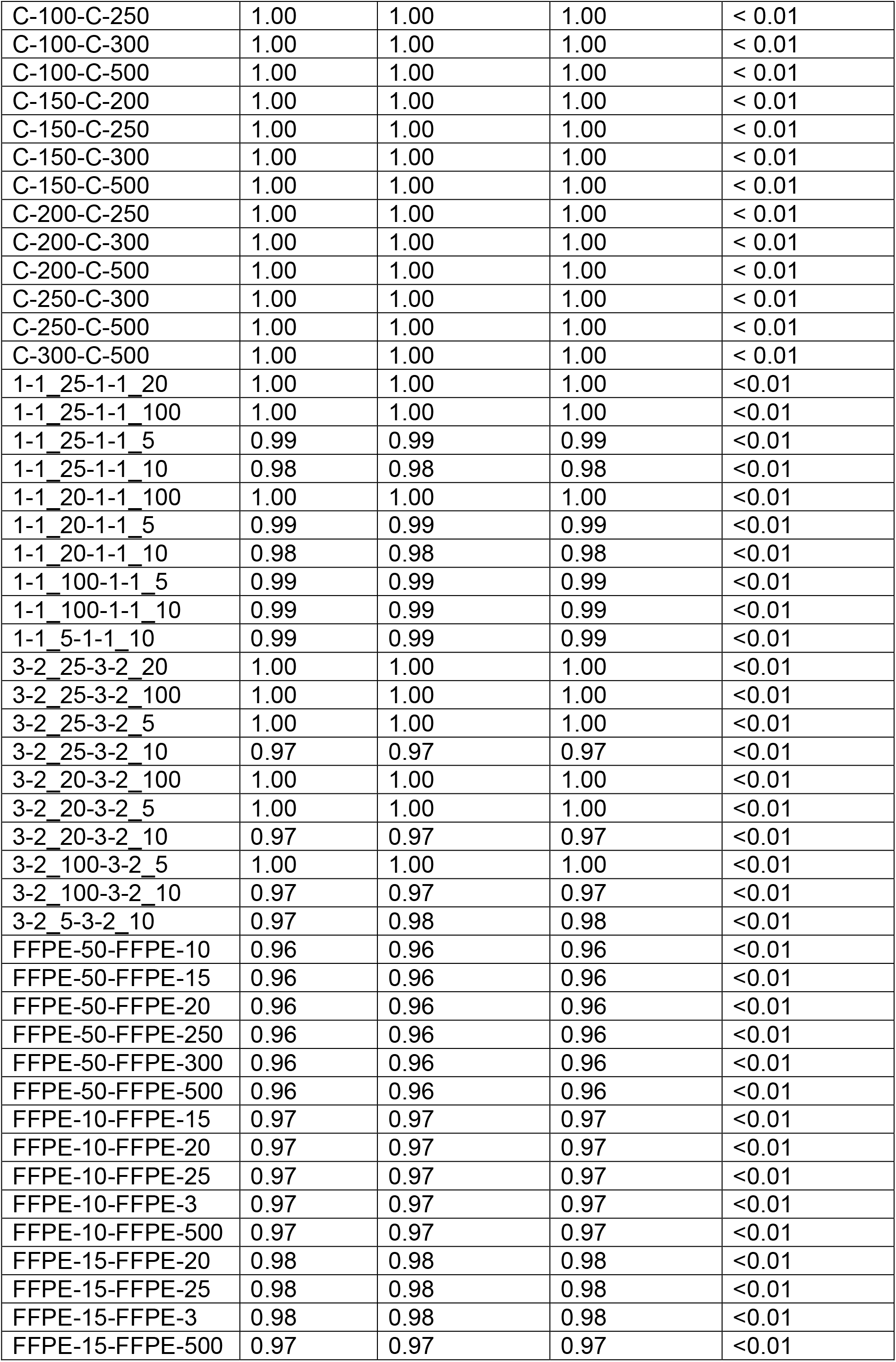
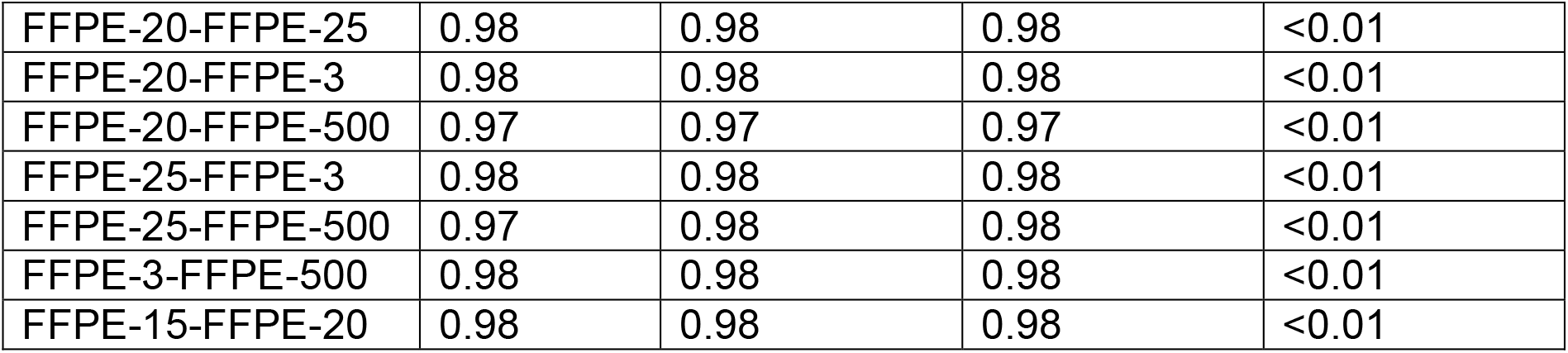
Correlation coefficients between all samples

**Figure 1:**
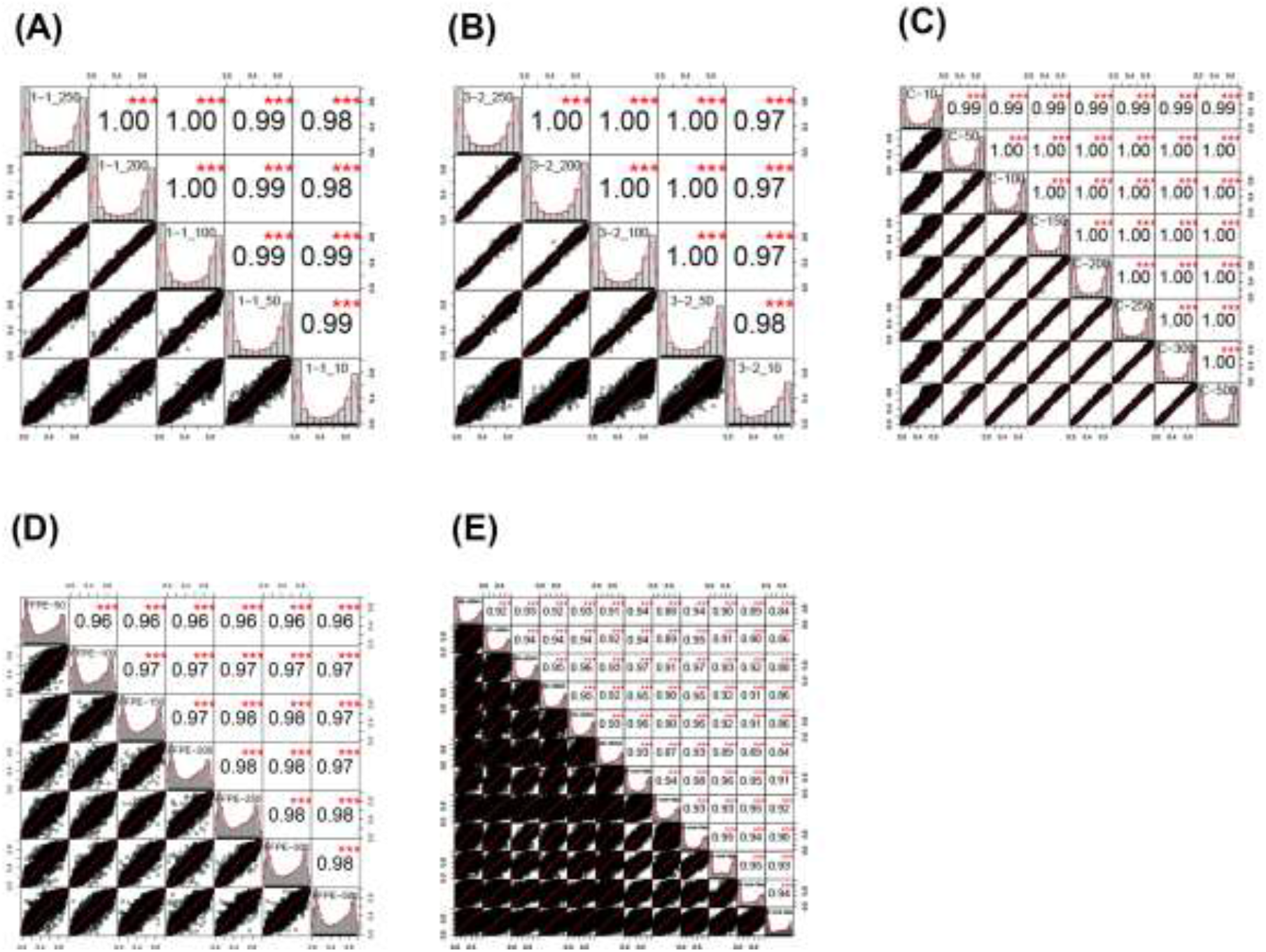
Pearson correlation plots of (A) Sample 1 (B) Sample 3 (C) Sample C (D) FFPE samples (E) ctDNA samples; Sample correlations are shown within box with labels down diagonal. Red stars represent statistical significance (*** = p < 0.001, ** = p < 0.01; * = p < 0.5). Black dot plots represent correlation between two samples.

#### Effect of input amount on results

In order to understand whether reducing sample amount impacted on results obtained through standard analysis, a differentially methylated position analysis was performed with the DMP analysis of R/ChAMP/Bioconductor with each sample randomly assigned to one of two groups, and each sample group analysed separately.

In the fresh blood derived sample group, for sample “1”, the DMP analysis demonstrated no significant differences between any of the groups comparing between each concentration, with the highest ranking probe cg17735539 having an d-Beta-meth of −0.12 an adjusted p-value of 0.99 and a Bayes Factor (B) of 1.96. For sample “3”, the DMP analysis demonstrated no significant differences between any of groups comparing between each concentration, with the highest ranking probe cg14640149 having a d-Beta-meth of −0.11, an adjusted p-value of 0.99 and a B of 0.41. For sample “C” the DMP analysis demonstrated no significant differences between groups, with the highest ranking probe cg18253591 having a d-Beta-meth of 0.04, an adjusted p-value of 0.43 and a B of 5.61.

In the FFPE sample group, for comparisons between groups, there was no significant difference with the highest ranking probe cg06871764 having a d-Beta-meth of 0.18, an adjusted p-value of 0.99 and a B of 2.77.

#### ctDNA analysis

In order to understand whether the observed low inputs would be suitable for detection of very fragmented, low input (typically 1-2ng) DNA, an experiment was carried out on six samples of circulating tumour DNA matched to their primary head and neck tumours of origin, sourced from fresh tissue.

All samples hybridised successfully to the arrays with very low input amounts when restored using the FFPE restore kit. One sample (288-cfDNA) had only 93% of probes that passed QC, possibly due to the increased fragmentation inherent to cfDNA. Median correlation between the tumour of origin and what was observed in the cfDNA samples was 0.92 (max 0.98, min 0.84). In the sample was the correlation was observed to be 0.84 (sample 288) this may be due to the poor quality of input sample.

In order to understand whether the observed differences were because of enrichment for “tumour” in the cfDNA samples over the contaminating normal stroma in the “tumour” samples which would exaggerate methylation differences, rather than contaminating genomic DNA, a standard methylation pipeline analysis using ChAMP was carried out. This was because if the cfDNA was mainly genomic DNA, it was expected that methylated regions typically seen in leucocytes (which are the predominant contributor to contaminating cfDNA) would be observed rather than those typical for HNSCC.

A differential methylation analysis revealed 19,014 differentially methylated positions between the control tumour samples and the circulating tumour DNA samples. The top ranked probe cg09568464 (chr19: 56904901) tagged a region within 200bp of the transcription start site of ZNF582 (deltaBeta=0.40, methcontrol=0.42, methcfdna=0.02, adjp=1.72×10-4, B=11.6). G:Profiler pathway analysis of the differentially methylated positions revealed the top enriched pathway was KEGG:04072 (Phosopholipase-D pathway, adj p value = 6.21×10-5) Several other tumoriogenic pathways of interest were also enriched including RAP1 signalling (padj=1.14×10-4), wnt signalling (padj=6.12×10-4), HPV infection (padj=9.56×10-3) and PIK3-Akt signalling (padj=0.02).

Analysis of differentially methylated regions revealed 410 DMRs with adjusted p values < 0.05. The top DMR was at chr6:29520965-29521803 (p<0.05) and tagged a region just upstream of UBD (ubiquitin-D) a key regulator of NF-kB. Pathway analysis utilising G:Profiler demonstrated two Kegg pathways that were enriched: KEGG:04080 (Neuroactive ligand receptor interaction, padj=6.73×10-4) and KEGG:04550 (Signalling pathways regulating pleuripotency of stem cells, padj=7.46×10-3).

#### DNA copy number calls

In order to ascertain whether copy number variation could be detected using this methodology in circulating tumour DNA, the copy prediction algorithm of conumee was used as per the vignette.

Generally, there was good agreement between copy number predictions for the reference sample and the ctDNA sample, with multiple copy number changes detected in both. There was dispersion of the copy number plot in the reference samples, most likely due to contaminating normal and the presence of multiple clones within the tumour sample. An example plot is shown in Figure 2, where the copy numbers between a fresh sample (top) and ctDNA derived sample from the sample patient (bottom) are compared. Copy number gains in CDKN2A and NOTCH are both observed in both samples, with high copy number seen in the ctDNA as compared to the reference sample. A similar pattern was seen in the FFPE derived samples (Figure 3) where increasing dispersion of copy number was seen as input DNA mass decreased.

**Figure 2:**
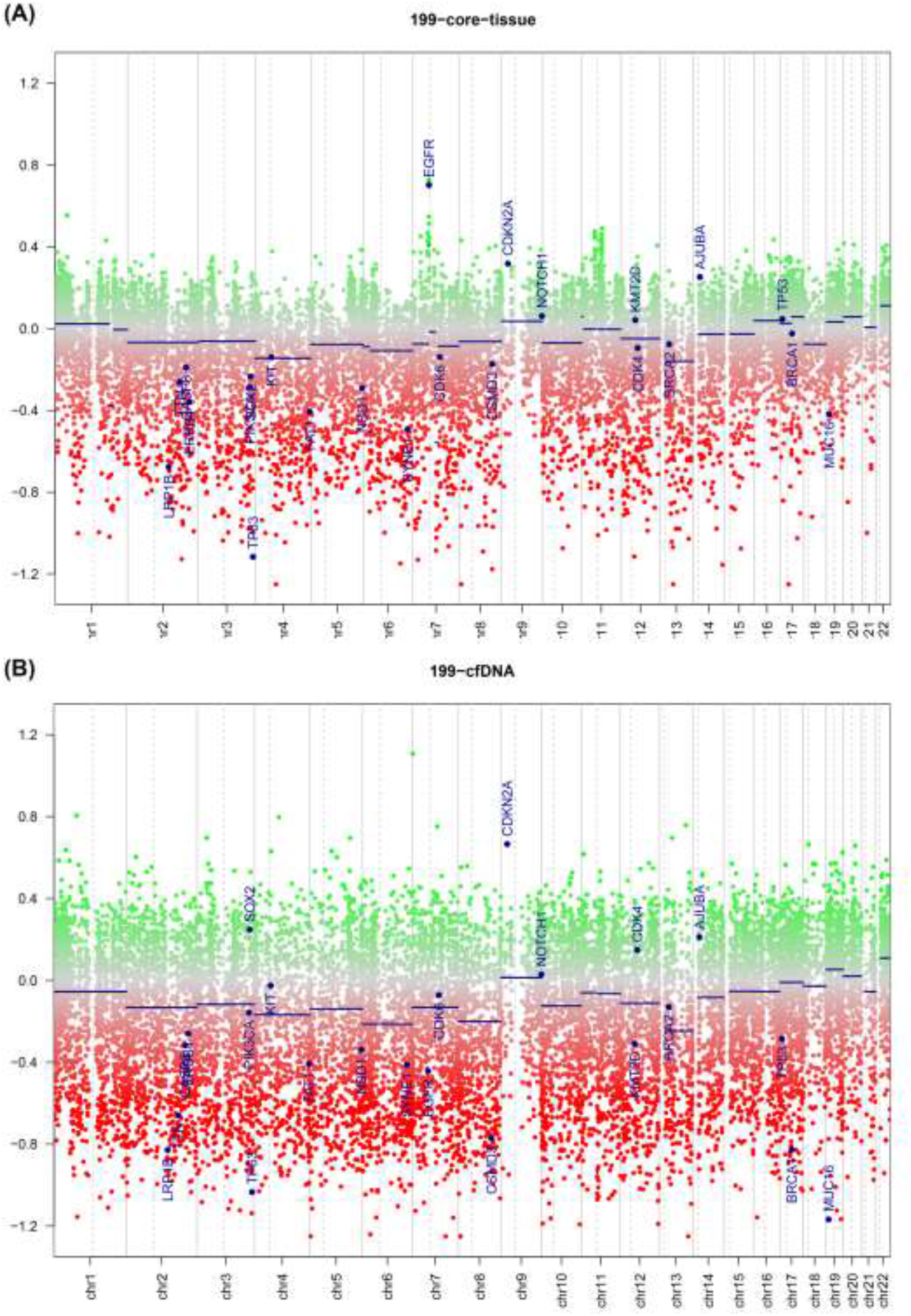
Copy number plot of reference fresh tissue (top) and ctDNA profile (bottom). Green dots = copy number gain; Red dots = copy number loss

**Figure 3:**
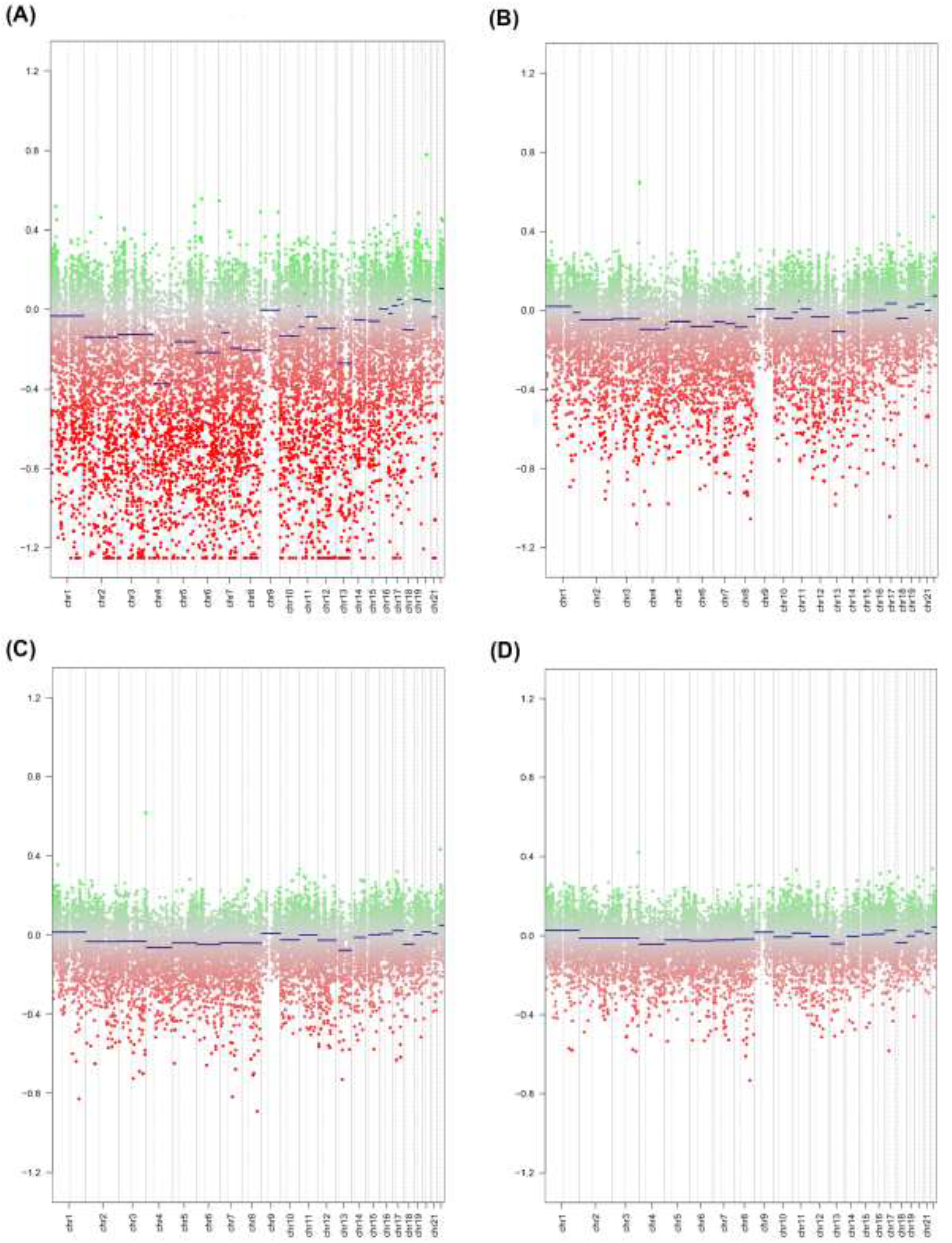
Copy number plots of a representative FFPE sample at (A) 10ng input (B) 50ng input (C) 100ng input (D) 250ng input. Green dots = copy number gain; Red dots = copy number loss

## DISCUSSION

Our results demonstrate that lower input to the Illumina Infinium assay as utilised in the MethylationEPIC and now unavailable Methylation 450K assay are feasible and provide reliable and reproducible results.

Because the core of the Infinium assay protocol is a whole genome amplification step, theoretical inputs into this kit could be as low as 1ng (15), and we have shown by our experiments on fresh blood derived samples, FFPE tissue and circulating tumour DNA that this target is achievable in future experiments. Methylation calls can be reliably and reproducibly carried out on ctDNA samples including detection of copy number variation with its origins in the base tumour.

This advance will allow access to previous inaccessible samples. Typical yields of ctDNA are between 1-10ng/ml of plasma (16) and so a typical blood draw will yield 4-40ng of ctDNA, of which ideally could now be divided between mutational and epigenetic assays. While previous strategies to overcome this low ctDNA input was to pool patient samples in risk defined groups (17) for exploratory purposes - this has no benefit for detecting prognostic signatures in individual patients for accurate risk stratification and/or treatment guidance. Also, biopsy samples of human disease states (e.g. cancer), especially when obtained from formalin-fixed, paraffin embedded specimens are typically very small and yield small amounts of DNA (18). These samples are now accessible to whole methylome analysis. For FFPE samples, we would recommend an input of at least 50ng of DNA, although smaller amounts are possible, with increasing inaccuracy, but if input DNA is not a limiting factor we would still suggest use at the recommended input into the assay.

Little, if any work has examined the whole epigenome of circulating tumour DNA. Several groups have demonstrated (19, 20) single gene techniques across a variety of cancer types. We have demonstrated that oncogenic methylation changes can be detected as well as copy number variants that are present in the original tumour. Recent work in DNA mutation burden from ctDNA samples suggest that the ctDNA seen represent the dominant metastatic clone in the cancer (21), and can be used for disease tracking (22). Several tumours demonstrate aberrant methylation as a key driver of their progression, including head and neck, brain and colorectal cancer. Epigenetic changes can be demonstrated from plasma samples, but these have been restricted to single gene promoters and thus make discovery work difficult (23, 24). Similarly, discovery work in human disease samples, e.g. in pre-treatment biopsies taken at endoscopy has also been limited due to the amount of input DNA present after extraction. This technique will allow access to these previously inaccessible disease states and allow temporal profiling of methylation i.e. pre-, during and post-treatment.

### CONCLUSION

In conclusion we have demonstrated that low input is possible into the Illumina HumanMethylation series of arrays, allowing access to new sample types that would have previously been inaccessible as well as detection of copy number variants and oncogenic methylation changes.

## METHODS

### Patient samples

All patient samples were obtained under either ethics for the S-CORT project (ref 15/EE/0241), the ethics for the Birmingham Human Biomaterials Resource Centre (ref 15/NW/0079), or the Accelerated (head and neck squamous cell carcinoma, HNSCC) tissue collection platform (REC ref: 16/NW/0265). In order to understand the effects of low input on FFPE derived samples (the SCORT cohort), DNA from the SCORT consortium was used. FFPE DNA samples were processed on Infinium® MethylationEPIC BeadChips in a single batch. Each sample had three varying DNA input amounts of 50 ng, 100 ng and 150 ng to give a 48 sample comparison. In order to understand the effects of DNA input on fresh DNA, blood derived samples three genomic control DNA samples (from a healthy donor) were processed on an Infinium® HumanMethylation 450k BeadChip (the Blood cohort). For this one sample input amount varied from 10 ng, 50 ng, 50ng, 100ng, 150ng, 200ng, 250ng, 300ng and 500 ng (labelled sample “C”). The other two DNA samples were processed using 10 ng, 50 ng, 100 ng, 200 ng and 250 ng input (labelled samples “1” and “3”. For the head and neck ctDNA samples (HNSCC samples), six HNSCC patient samples were obtained pre-treatment from the Accelerated tissue collection platform – fresh tissue and blood (collected in Streck blood tubes) - and processed on an Infinium® HumanMethylation 450k BeadChip.

For the Infinium® MethylationEPIC BeadChip and Infinium® HumanMethylation 450k BeadChip the Illumina recommended input amount is ≥ 250 ng.

### DNA extraction

Human FFPE DNA was extracted from between 6 and 9, 5-micron FFPE colorectal tumour blocks using the QiaAmp Micro Kit (Qiagen). A marked up H&E stained section was used to guide macrodissection of tumour tissue. An assessment of DNA quality and quantity was made initially using a Nanodrop 1000 3.3.0, then the quantity of double stranded DNA was determined using the Quant-iT Picogreen ds DNA Assay Kit (Life technologies). Sample concentrations were checked and normalised to 20 ng (6 samples) and 250 ng (6 samples).

Genomic DNA was extracted from HNSCC fresh tissue biopsies using the DNeasy blood and tissue kit (Qiagen). Cell-free DNA was extracted from 2ml of plasma from HNSCC blood samples using the QIAamp circulating nucleic acid kit (Qiagen). Sample concentrations were checked and normalised to 20 ng (6 blood samples) and 250 ng (6 tissue samples).

For the blood control samples, a Promega Maxwell RSC instrument (AS4500) was used with the Maxwell RSC Whole Blood DNA Kit (AS1520). This is a semi-automated DNA extraction method which utilises paramagnetic particles in pre-loaded cartridges to bind DNA and elute into 20 ul volumes. The purified DNA was quantified using the Qubit 2.0 Fluorometric Quantitation instrument using the dsDNA BR (broad range) assay.

### FFPE QC Assay

To determine FFPE DNA suitability for the Infinium HD FFPE methylation assay the quality was tested in duplicate by real-time PCR following the Illumina FFPE QC protocol (Part # 15020981 Rev. C). Amplification of FFPE sample DNA was compared with the amplification of a Quality Control Template (QCT). The real-time PCR threshold cycle (Ct) was averaged and a DCt for each sample was calculated (DCt = CtFFPE - CtQCT). An FFPE DNA sample was deemed suitable if the DCt was <5.

#### Bisulphite Conversion & Restoration of DNA

The FFPE DNA was bisulphite converted using the EZ DNA Methylation Kit (cat# D5002, Zymo Research) following the manufacturer’s instructions appropriate for the Illumina HD FFPE assay. A total of 8 µl of DNA was eluted and taken through to the next stage FFPE DNA Restoration. For non-FFPE DNA 4 ul was taken through directly to the Infinium HD FFPE Methylation Assay. FFPE DNA restoration was achieved by using the Infinium HD FFPE restore protocol. This restores degraded FFPE DNA to a state that is amplifiable. All eluted restored DNA (approx. 8 µl) was taken through to the Infinium HD FFPE methylation assay.

#### Infinium HD FFPE Methylation Assay

All DNA samples were processed following the Infinium HD FFPE Methylation Assay, Manual Protocol. For the Infinium® MethylationEPIC BeadChips the appropriate RA1 resuspension and hybridising volume were used as provided in the Infinium HD Methylation Assay Manual Protocol. The Illumina iScan was used to scan the arrays recording high resolution images of light emitted by the excited fluorophores at each CpG site on the arrays. The raw intensity data from these images was stored as .idat files and used for analysis. Illumina GenomeStudio v2011.1 was used to process the .idat files. Sample dependent and independent controls were analysed to ensure the assay procedure had been successful. The % of CpG detection was determined at a confidence level of p=0.05.

All samples passed QC. The analysis had no sample groupings or normalisation, and the background was subtracted.

### Downstream analysis

All Idat files were imported into R/Bioconductor and analysed using ChAMP (25) pipeline up to DMP/DMR processing. ChAMP is a R/Bioconductor pipeline that reads raw data from Illumina iDat files, carries out filtering and quality control and can carry out all downstream processing. For intra-group comparisons, the probes for each concentration level were compared to every other concentration using multilevel regression. For copy number calling, the conumee algorithm (26) was used. For between sample correlations the *cor* command of the *psych* module from CRAN (https://cran.r-project.org/web/packages/psych/index.html) was used. For pathway analysis, top ranked differentially expressed DMP/DMR were exported to GProfiler (27).

## Data Availability

Data will be made available on acceptance of manuscript

## DECLARATIONS

### Ethical approval

All patient samples were obtained under either ethics for the S-CORT project (ref 15/EE/0241),the ethics for the Birmingham Human Biomaterials Resource Centre (ref 15/NW/0079), or the Accelerated (head and neck squamous cell carcinoma, HNSCC) tissue collection platform (REC ref: 16/NW/0265)

### Availability of data and materials

All data will be made available on GEO on publication of manuscript

### Contributorship

Manuscript: ADB, KP; Molecular analysis: CW, AB; Sample acquisition and patient cohort: All authors; Bioinformatics: ADB, AB, ED.

